# Cancer risks for *MSH6* pathogenic variant carriers

**DOI:** 10.1101/2025.02.15.25322330

**Authors:** A.S. van der Werf – ‘t Lam, J.G. Dowty, M. Italia, A.C. Bakker, F. Koops, F. Bleeker, E. Gomez – Garcia, L.P. van Hest, H.J.P. Gille, M.C. Cornips, M.M. de Jong, T.G.W. Letteboer, F.A.M. Duijkers, A. Wagner, E.L. Eikenboom, C.J. van Asperen, S.W. Bajwa – ten Broeke, A.K. Win, M.A. Jenkins, M. Nielsen

## Abstract

**Introduction:** Lynch syndrome (LS) is a hereditary cancer syndrome caused by (likely) pathogenic variants (LP/P) in DNA mismatch repair genes, including *MSH6*. It is associated with elevated lifetime risks for colorectal cancer (CRC), endometrial cancer (EC), and other malignancies. However, cancer risks specific to *MSH6*-associated LS, particularly for non-colorectal cancers, remain poorly defined. This study aims to provide refined cancer risk estimates for individuals with *MSH6* LP/P.

**Methods:** We conducted a retrospective cohort study of 360 families with 1117 known *MSH6* LP/P carriers identified in the Netherlands between 1995 and 2020. Pedigree data were collected from multiple clinical centers, and cancer diagnoses were confirmed through medical records. Age- and sex-specific hazard ratios (HRs) and cumulative risks (CRs) were estimated using segregation analysis, appropriately adjusted for ascertainment.

**Results:** CR by age 80 for *MSH6* LP/P carriers were 36% in males (95% CI:25–48%) and 21% in females (95% CI 13–32%) for CRC, and 23% in females (95% CI:15–43%) for EC. Elevated risks were observed for ovarian cancer (OC) (6.4%, 95% CI:3–14.8%; HR 5.58, p=0.00037), urinary tract cancers (10.1% in males, 4.1% in females; HR 2.52, p=0.012), and biliary tract cancers (4.9% in males, 4.2% in females; HR 2.76, p=0.031). No increased risks were identified for prostate or breast cancer.

**Conclusion:** This study refines cancer risk estimates for MSH6 LP/P carriers, suggesting the need for delayed CRC screening in males and females and proactive discussions regarding prophylactic surgery for females to address elevated risks for EC and OC.

## Introduction

Lynch syndrome (LS) is a hereditary condition characterized by a heterozygous (likely) pathogenic variant (LP/P) in one of the mismatch repair (MMR) genes: *MLH1, MSH2, MSH6, PMS2*, or *EPCAM* deletions [1]. The estimated carrier frequency of LS is approximately 1 in 279 individuals [2]. LS is primarily linked to a significantly increased lifetime risk (LTR) of colorectal cancer (CRC) and endometrial cancer (EC). However, studies have also reported an elevated LTR for other extracolonic cancers, including ovarian, stomach, small bowel, bile duct, and upper urinary tract cancers [3]. These cancer risks vary based on the specific MMR gene involved [3, 4], but for CRC, also across different geographic regions, likely due to genetic and environmental risk factors [5]. The implementation of universal tumor screening for mismatch repair deficiency and the increasing use of multigene panel testing further underscores the importance of these variations.

Individuals diagnosed with LS can benefit from regular cancer screening and risk-reducing surgeries. Frequent colonoscopies have been proven effective in reducing the risk of CRC [6-9], while prophylactic hysterectomy and salpingo-oophorectomy can prevent LS-associated EC and ovarian cancer [10]. However, accurate risk estimates, especially for *MSH6-*associated LS, remain uncertain, complicating patient surveillance and counseling.

Previous larger studies have reported varying cancer risks for *MSH6*-associated LS, often with wide confidence intervals. For example, the LTR reported for CRC ranges from 10.0% to 20.3%, and for EC, it varies between 16% and 41.1% [3, 11, 12]. Although these risks are consistently higher than those in the general population [13], the LTR for other cancers is less clear. For instance, the reported LTR for ovarian cancer (OC) ranges from population risk [11] to 10.8% (95% CI: 3.7-32.9%) [3], and similarly, conflicting LTRs have been reported for breast cancer[3, 14-16].

Therefore, the aim of this study is to assemble a large cohort of families with an *MSH6* LP/P variant to derive more accurate cancer risk estimates for this specific subgroup of LS patients. By achieving this, we aim to enhance the current guidelines and improve surveillance and management strategies for individuals with *MSH6*-associated LS.

## Methods

### Data collection

Family pedigrees were collected of all known *MSH6* families in whom a (likely) pathogenic variant was detected from 1995 until January 2020 in the collaborating centers. The minority of these *MSH6* mutation carriers were identified because of universal testing that was conducted irrespective of family history, and therefore analyzed as population-based ascertainment. The remainder were assumed to have been identified due to testing based on presence of cancer family history, and therefore analyzed as clinic-based ascertainment. These families were collected in one of the following clinical genetic centra in the Netherlands: Dutch Cancer Institute, Maastricht University Medical Center, Amsterdam UMC, location Vrije Universiteit Amsterdam and location University of Amsterdam, University Medical Center Groningen, University Medical Center Utrecht, Erasmus Medical Center and Leiden University Medical Center. Beforementioned contributing clinical genetics departments provided coded family pedigrees, including the following data: sex, carrier status of *MSH6* variants, and any other variants (i.e., a carrier, non-carrier, or unknown/untested), age at last contact or death. Where applicable, the age of cancer diagnosis (anatomical site and age at diagnosis), age of first polypectomy or start colonic screening, age of hysterectomy, age of ovariectomy, or age of mastectomy were provided. Data quality and consistency were checked by a data parser, and incomplete or inconsistent data was redressed. Counseled patients were contacted through the clinical genetics departments for informed consent to request their pathology reports or medical records to confirm a cancer diagnosis, polypectomies, and (risk reduding) hysterectomies where possible.

This study was approved by the Medical Ethical Committee of Leiden, The Hague, Delft (protocol P17.098).

### Variant analysis and clinical variant classification

Probands were tested based on immunohistochemistry and/or microsatellite instability results, while relatives were screened for the known variant in the respective family. Variants were classified for pathogenicity according to the InSiGHT Variant Interpretation Committee Mismatch Repair Gene Variant Classification Criteria and published on the LOVD [17].

### Statistical analysis

Cancer risks for carriers of *MSH6* LP/P variants were estimated using segregation analysis, since this method can incorporate adjustments for complex ascertainment schemes and can use known genetic laws to account for statistical dependence between family members. Full mathematical details of a similar approach can be found in a previous report [18].

Estimates were adjusted for the ascertainment of families using either a retrospective likelihood or an ascertainment-corrected joint likelihood [19-22] in which each clinic-based pedigree’s data was conditioned on the proband’s genotype and the ages and affected statuses of all family members, and each population-based pedigree’s data was conditioned on the genotype, ages and affected statuses of the proband. This approach gives risk estimates that are unbiased by ascertainment, even when the exact ascertainment scheme for clinic-based families cannot be modeled, as long as their ascertainment only depends on the families’ phenotypes [20].

Cancer incidences for non-carriers were set equal to the sex- and age-specific population incidences from the IKNL database, averaged over the years from 2015 to 2019 [13]. For carriers, cancer incidences were the product of an age- and sex-specific hazard ratio, which we estimated, multiplied by the corresponding non-carrier incidence.

Hazard ratios were modeled as a piecewise linear function of age that was constant before age 40 and after age 60 and linear between these ages. We estimated hazard ratios for CRC and EC simultaneously; these estimates were then fixed, and the hazard ratios for other the cancer types were estimated one at a time.

We adopted a population allele frequency of 1/1516, as reported by Win et al. [2], though our results would likely be the same for all plausible allele frequencies. Additionally, we introduced a polygenic model to account for familial aggregation of colorectal cancer, incorporating factors beyond MSH6 variants, with a polygenic standard deviation of 1.682 based on European estimates from the IMRC study [5].

We excluded 7 families without any age information, and we excluded third- or higher-degree relatives of the proband from each family. Individuals were censored at the earliest of death, last follow-up, age 80, polypectomy (for CRC only) or hysterectomy (for EC only). Participants with missing sex (199 individuals) were censored at birth, i.e. treated as unaffected and aged 0 years.

Cancer affected individuals with missing ages at diagnosis (56 CRC, 16 EC, 188 other cancers) were given imputed ages equal to the median age at diagnosis for their sex and specific cancer type in the Netherlands 2015-2019. Unaffected individuals with missing ages (4945 individuals) were given imputed ages equal to (in order of decreasing preference) either the average age of their full siblings, their half-siblings, their spouses, their parents minus 25 years, or their offspring plus 25 years. These imputation rules were then iterated until no missing ages remained.

All statistical analyses were conducted in R version 4.2.0, utilizing the *clipp* package to calculate pedigree likelihoods and maximizing the likelihoods using the *mle* function, as in Ryan et al. [23]. Statistical significance was assessed with the likelihood ratio test.

## Results

### Overview

A total of 360 families were included accountable for 2582 first-degree and 5298 second-degree relatives of whom 757 were confirmed carriers and 821 were confirmed non-carriers of a LP/P *MSH6* variant (see also Table 1). Table 2 displays the number of cases and mean ages at diagnosis for each cancer site among first- and second-degree relatives.

**Table 1.** Study data set description. Abbreviations: FDR = first-degree relative; SDR = second-degree relative

| Individuals | No. of relatives |  |  |
| --- | --- | --- | --- |
|  | Total | Male | Female |
| **Probands = (no. of families)** | 360 | 139 | 221 |
| - FDR | 2582 | 1292 | 1290 |
| - SDR | 5298 | 2633 | 2665 |
| **Confirmed *MSH6* variant carriers** |  |  |  |
| - FDR | 476 | 230 | 246 |
| - SDR | 281 | 112 | 169 |
| **Confirmed *MSH6* non carriers** |  |  |  |
| - FDR | 493 | 209 | 284 |
| - SDR | 328 | 146 | 182 |

**Table 2.** No. and mean age at diagnosis of each cancer site in the FDRs and SDRs of probands. Abbreviations: FDR = first-degree relative; SDR = second-degree relative; IQR = interquartile range

| Anatomical site | FDR (n = [2582]) |  | SDR (n = [5298]) |  |
| --- | --- | --- | --- | --- |
|  | No. | Median (IQR)<br>age at diagnosis<br>(years) | No. | Median (IQR)<br>age at diagnosis<br>(years) |
| **colorectum** | 257 | 63 (18) | 226 | 64 (17) |
| **endometrium** | 113 | 58 (12) | 73 | 58 (13) |
| **small bowel** | 5 | 62 (19) | 1 | 62 (0) |
| **biliary tract** | 17 | 72 (6) | 43 | 72 (5) |
| **ovary** | 20 | 60 (19) | 15 | 52 (24) |
| **prostate** | 31 | 71 (5) | 41 | 71 (0) |
| **urinary tract** | 39 | 71 (12) | 30 | 71 (2) |
| **brain** | 24 | 64 (27) | 22 | 64 (8) |
| **breast** | 54 | 60 (13) | 92 | 62 (13) |

Table 2 shows the number and mean age at diagnosis (with standard deviation) for various cancer sites among first- and second-degree relatives of *MSH6* variant carriers.

### Cancer risks

An overview of cancer risks are depicted in Table 3 and 4 and in Figure 1 and 2.

**Table 3.**
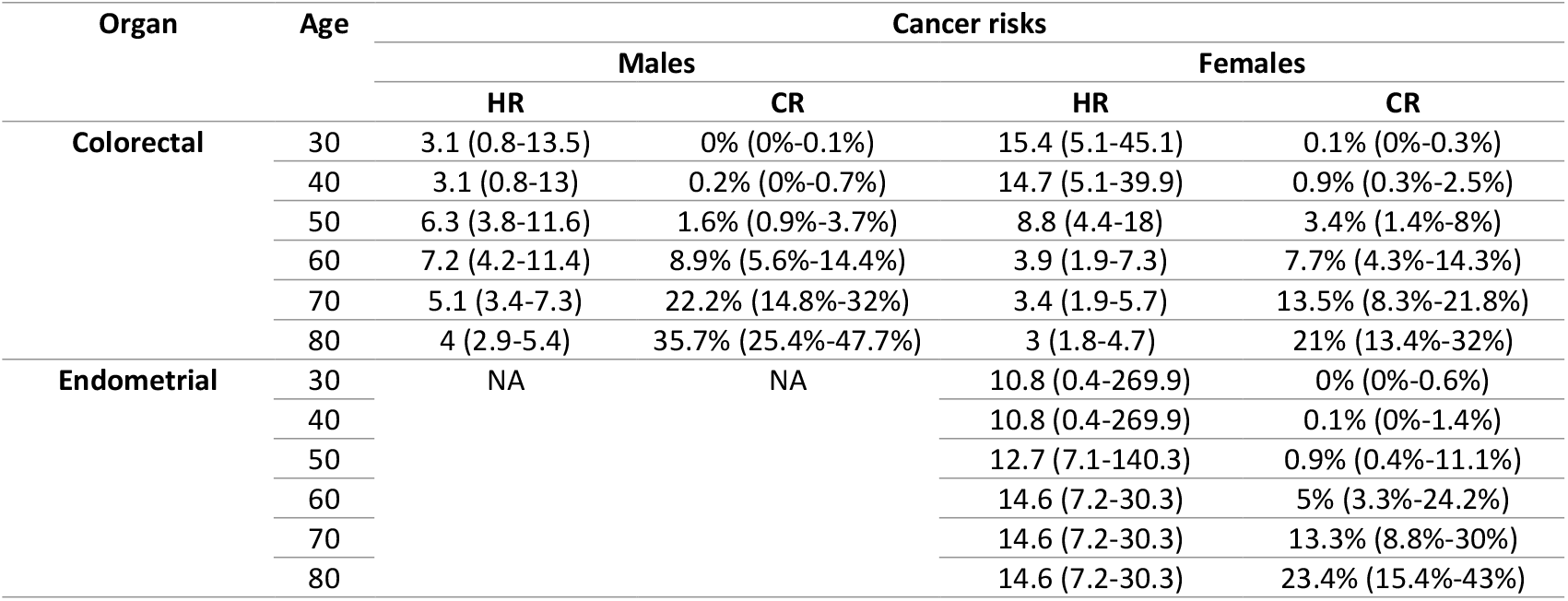
Cancer risks subdivided for men and women. HR = hazard ratio; CR = cumulative risk; NA = not applicable;

**Table 4.** extra colonic cancer risks, excluding endometrial cancer, subdivided for men and women. HR = hazard ratio; CR = cumulative risk; NA = not applicable;

| Organ | Age | Male | Female |
| --- | --- | --- | --- |
|  |  | CR % (95% CI) | CR % (95% CI) |
| Ovarian | 30 | NA | 0.4 (0.2-1.2) |
|  | 40 |  | 1.5 (0.5-4) |
|  | 50 |  | 3.5 (1.3-9.1) |
|  | 60 |  | 5.1 (2-12.8) |
|  | 70 |  | 5.6 (2.4-13.6) |
|  | 80 |  | 6.4 (3-14.8) |
| Biliary tract including: stomach, bile duct, gallbladder, and pancreas | 30 | 0.0 (0.0 - 0.0) | 0.0 (0.0 - 0.0) |
|  | 40 | 0.0 (0-0.1) | 0.0 (0-0.1) |
|  | 50 | 0.2 (0.1-0.3) | 0.1 (0.1-0.3) |
|  | 60 | 0.7 (0.3-1.6) | 0.6 (0.3-1.4) |
|  | 70 | 2.2 (1-4.8) | 1.9 (0.8-4.3) |
|  | 80 | 4.9 (2.2-10.8) | 4.2 (1.8 - 9.4) |
| Urogenital tract including: ureter, kidney urinary bladder | 30 | 0 (0-0.1) | 0 (0-0.1) |
|  | 40 | 0.1 (0.1-0.3) | 0.1 (0.1-0.2) |
|  | 50 | 0.5 (0.3 - 1) | 0.3 (0.1-0.5) |
|  | 60 | 1.7 (0.9-3.4) | 0.8 (0.4-1.7) |
|  | 70 | 4.7 (2.4-9.0) | 2.0 (1.0 - 4.0) |
|  | 80 | 10.1 (5.2-18.8) | 4.1 (2.1 - 8.1) |
| Small bowel | 30 | 0 (0-0.1) | 0 (0-0.1) |
|  | 40 | 0 (0-0.1) | 0 (0-0.1) |
|  | 50 | 0 (0-0.1) | 0 (0-0.1) |
|  | 60 | 0 (0-0.3) | 0 (0-0.2) |
|  | 70 | 0.1 (0-1.2) | 0 (0-0.7) |
|  | 80 | 0.2 (0-2.8) | 0.1 (0.1-1.9) |
| Prostate | 30 | 0 (0-0) | NA |
|  | 40 | 0 (0-0) |  |
|  | 50 | 0.1 (0.1-0.3) |  |
|  | 60 | 1.7 (0.8-3.4) |  |
|  | 70 | 7.8 (3.8-15.1) |  |
|  | 80 | 16.6 (8.4-30.6) |  |
| Brain | 30 | 0.1 (0-0.4) | 0 (0-0.2) |
|  | 40 | 0.2 (0.1-0.8) | 0.1 (0-0.4) |
|  | 50 | 0.4 (0.1-1.5) | 0.2 (0.1-0.9) |
|  | 60 | 0.7 (0.2-2.8) | 0.4 (0.1-1.7) |
|  | 70 | 1.2 (0.3-4.7) | 0.7 (0.2-2.8) |
|  | 80 | 1.8 (0.5-7.2) | 1.1 (0.3-4.4) |
| Breast | 30 | NA | 0.1 (0.1-0.2) |
|  | 40 |  | 0.7 (0.4-1.4) |
|  | 50 |  | 2.5 (1.4-4.6) |
|  | 60 |  | 5 (2.8-9) |
|  | 70 |  | 8 (4.5-14.3) |
|  | 80 |  | 10.8 (6.2-19) |

**Figure 1.**
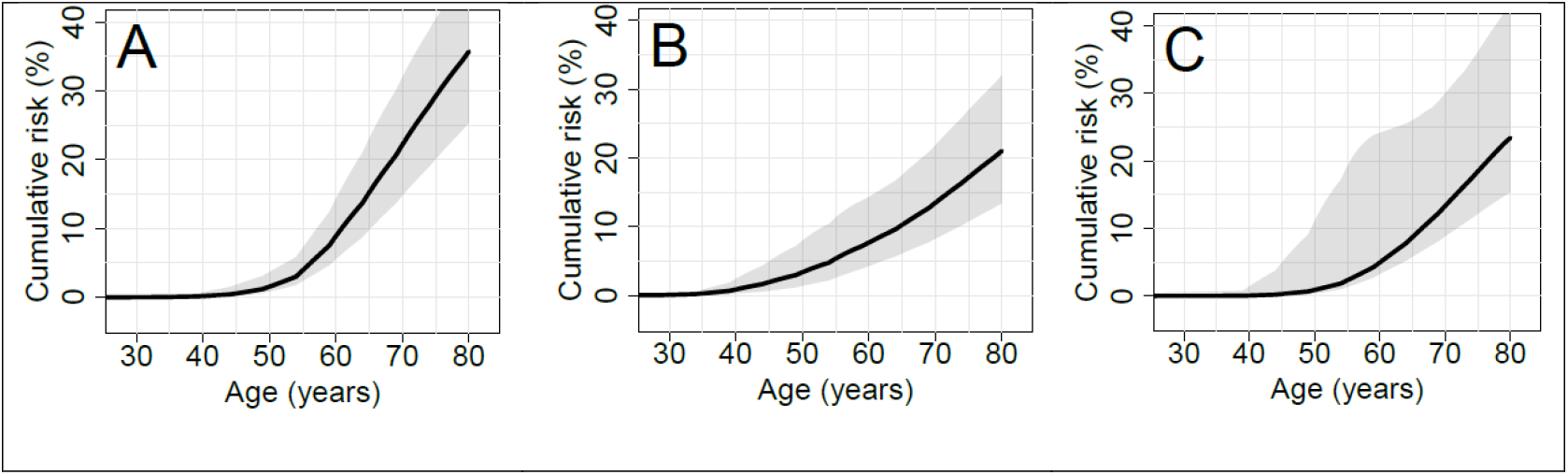
Cumulative cancer risks A= colorectal cancer risk for males, B= colorectal cancer risk for females, C= endometrial cancer

**Figure 2.**
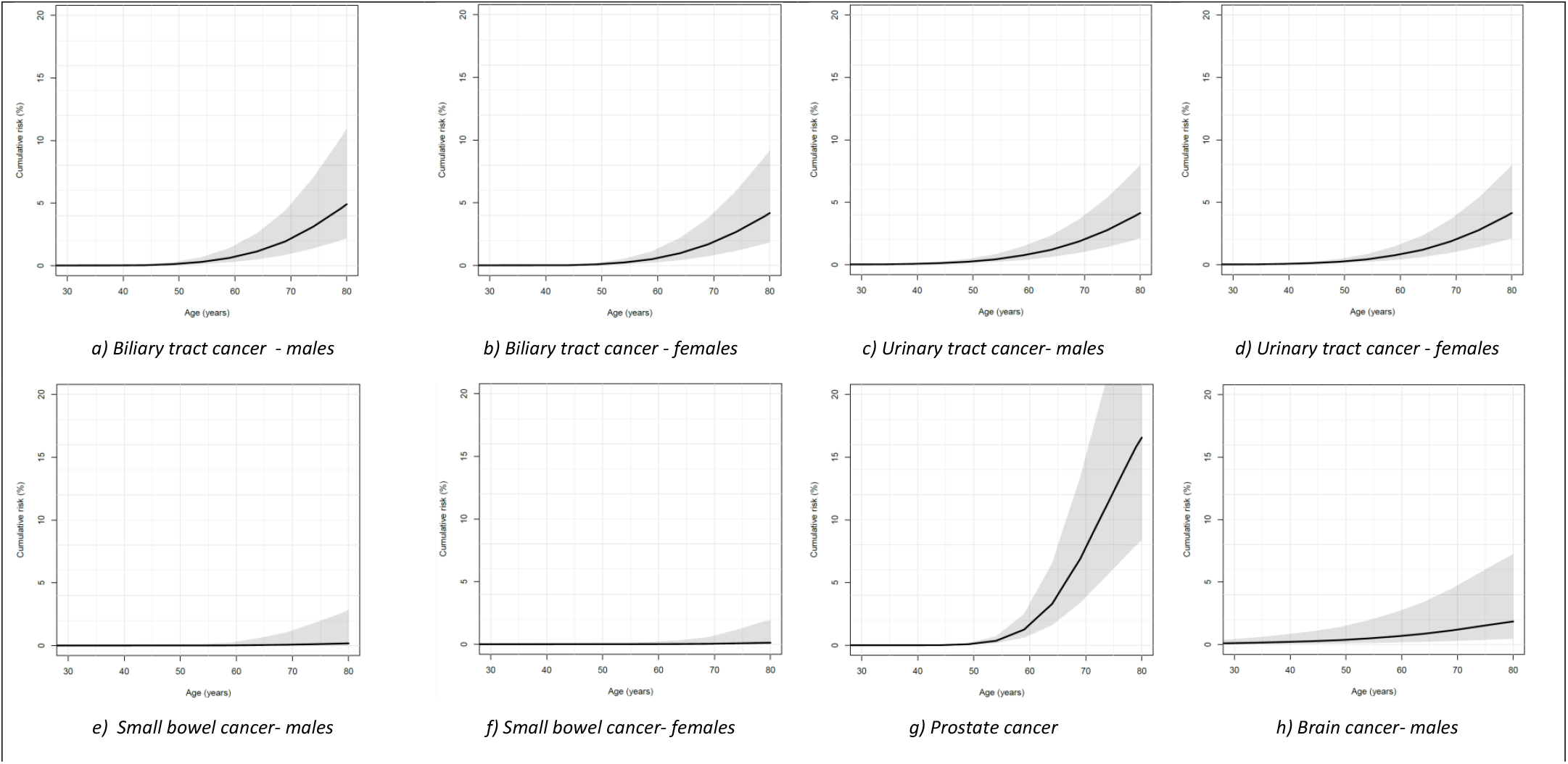
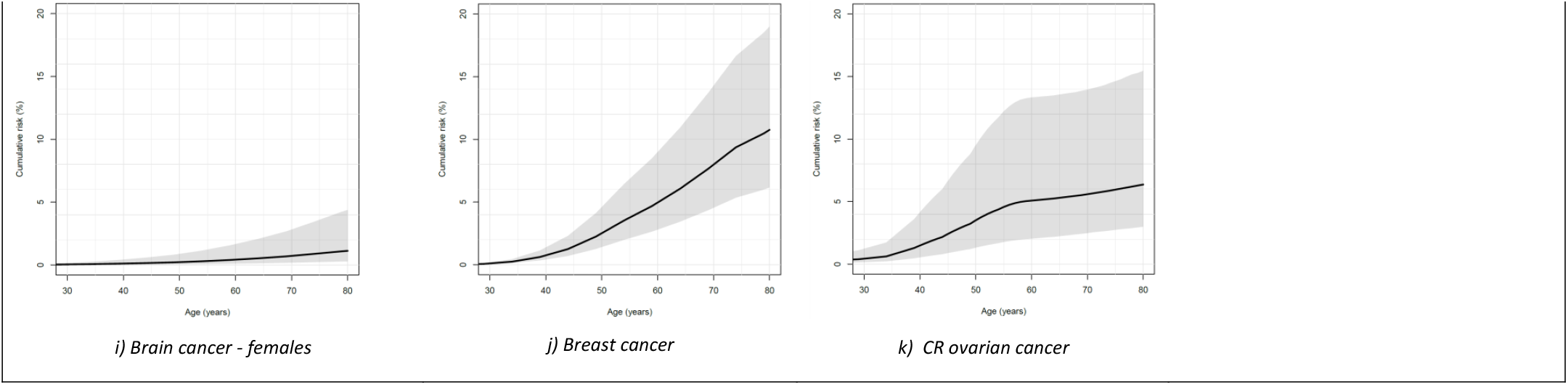
Cumulative cancer risks in %

#### Colorectal cancer

Carriers of a *MSH6* LP/P variant have an elevated risk of developing CRC, with hazard ratios varying by age and sex. Specifically, the HRs are 3.1 (0.8-13), resulting in a CR of 0.2% (95% CI: 0.2% - 0.7%) for males under 40 years, and 5.1 (3.4-7.3), resulting in a CR of 22.2% (95% CI: 14.8%-32.0%) for males over 70 years. For females, the HRs are 14.7 (5.1-39.9), resulting in a CR of 0.9% (95% CI: 0.3%-2.5%) at 40 years, and 3.4 (1.9-5.7), resulting in a CR of 13.5% (95% CI: 8.3%-21.8%) over 70 years. The estimated CR of developing CRC by age 80 for *MSH6* LP/P variant carriers is approximately 35.7% (95% CI: 25.4%-47.7%) for males, and 21% (95% CI: 13.4%-32%) for females, compared to 6.6% and 4.7% in the general population, respectively (See Figure 1).

#### Gynaecologic cancers

The CR at the age of 80 years for endometrial cancer is 23.4% (95%CI: 15.4%-43%), and 6.4% (95%CI: 3%-14.8%)for ovarian cancer (see also Figure 1). Compared to the population risk, both cancer risks are increased.

#### Other cancers

The cumulative risk (CR) of urinary tract cancer at age 80 is 10.1% (5.2%–18.8%) for males and 4.1% (2.1%–8.1%) for females. For biliary tract cancer, the CR is 4.9% (2.2%–10.8%) for men at age 80 and 4.2% (1.8%–9.4%) for women. See also Table 4 for further details. Additionally, Table 5 presents the hazard ratios (HR) for small bowel, biliary tract, ovarian, prostate, urinary tract, brain, and breast cancers. There was no evidence that HRs depended on sex (all p > 0.7) or age (all p >= 0.2), with the sole exception that ovarian cancer HRs depended on age (p < 0.001), with estimated HRs of 38.8 (13.9-108) at age 40 and 1.54 (0.43-5.53) at age 60.

**Table 5.**
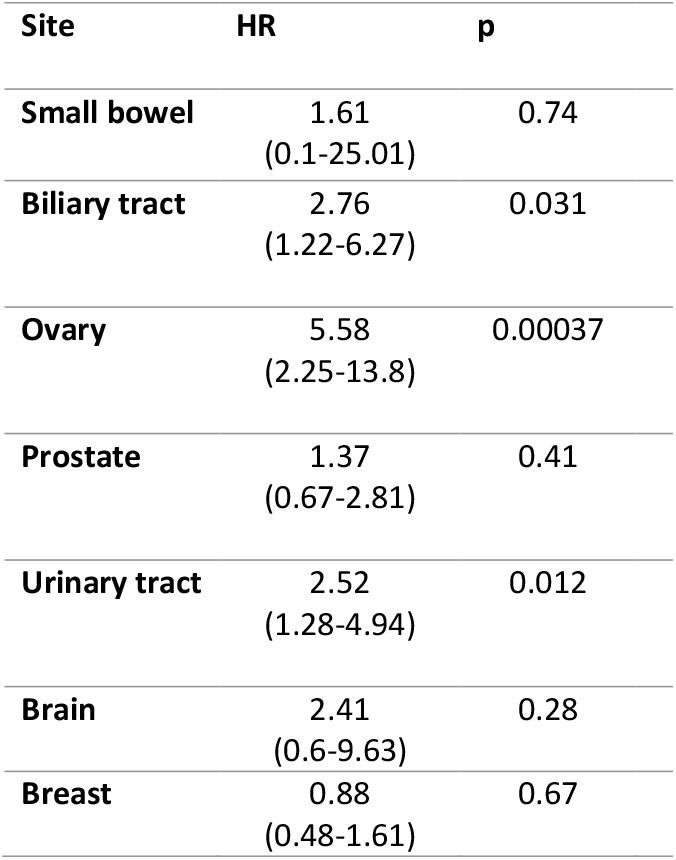
HR extracolonic cancers.

| Site | HR | p |
| --- | --- | --- |
| Small bowel | 1.61<br>(0.1-25.01) | 0.74 |
| Biliary tract | 2.76<br>(1.22-6.27) | 0.031 |
| Ovary | 5.58<br>(2.25-13.8) | 0.00037 |
| Prostate | 1.37<br>(0.67-2.81) | 0.41 |
| Urinary tract | 2.52<br>(1.28-4.94) | 0.012 |
| Brain | 2.41<br>(0.6-9.63) | 0.28 |
| Breast | 0.88<br>(0.48-1.61) | 0.67 |

## Discussion

This study aimed to provide more accurate cancer risk estimates for *MSH6* variant carriers by assembling a large cohort of families with *MSH6* LP/P variants. Our findings highlight the increased cancer for risk for EC and CRC in these individuals compared to the general Dutch population, with variations depending on age and sex. These insights are crucial for refining surveillance and management strategies for LS patients with *MSH6* LP/P variants.

The variability in cancer risk estimates reported in previous studies [3, 11, 12, 24] reflects the complexities in assessing cancer risk for *MSH6*–associated LS. Our study aligns with earlier findings but provides a more nuanced understanding of how risk fluctuates with age and sex for the Dutch population. These insights are crucial for refining clinical guidelines and improving patient counselling. A more tailored approach based on these age and sex variations could lead to better risk stratification, allowing clinicians to offer more precise advice regarding surveillance and preventive measures. This granularity in risk assessment is particularly useful for making informed decisions about when to initiate screening, as well as for determining the appropriate intervals between screenings.

### Colorectal cancer risks and screening

The current NCCN Guidelines [25] recommend initiating CRC surveillance with colonoscopy at age 25 for *MSH6* carriers. However, our data suggest that a later start, potentially at age 30 or 35 – or even later – could be appropriate, particularly for males, given the relatively low CRC risk in individuals under 50. The hazard ratios (HR) and cumulative risk (CR) observed in our cohort imply that postponing colonoscopy could reduce unnecessary procedures and the associated medical and psychological burden without significantly increasing cancer risk. This approach aligns with evolving perspectives on more individualized surveillance strategies.

Additionally, for those starting colonoscopies later, adjusting the screening interval may help balance effectiveness and burden. We propose a more gradual approach, such as beginning screening at age 30, followed by a second colonoscopy at age 35, and then transitioning to biennial screening from age 40 onward. This strategy aims to reduce the frequency of unnecessary procedures during younger ages while maintaining effective cancer prevention.

When comparing these findings to other studies, the recent analysis by Dominguez-Valentin et al. (2023) [26] reported notably higher CRC risks for *MSH6* carriers than seen in prior studies [3, 27, 28] Possible explanations for this discrepancy include differences in population sampling, study design, and ascertainment biases. Further investigation is warranted to reconcile these findings.

### Endometrial cancer risks and risk reducing surgery

For women with pathogenic *MSH6* variants, the decision to consider risk reducing hysterectomy and oophorectomy is a crucial consideration due to the substantial risk of endometrial cancer (up to 23.4% by age 80) and moderate risk of ovarian cancer (up to 6.4% by age 80). The NCCN guidelines recommend discussing these risk reducing surgeries after childbearing, generally around age 35-40 [25]. Similarly, the Manchester International Consensus Group advises risk-reducing surgery at age 40 for *MSH6* carriers [29]. However, the lack of stratified studies specific to LS-associated endometrial and ovarian cancers complicates individualized recommendations.

Our findings affirm the rationale for early discussion from age 40-45 of risk reducing surgery, particularly for women who have completed family planning. The high cumulative cancer risks identified in our study affirm the need for a proactive individualized approach, taking into account cancer risk, reproductive plans, and patient preferences. Moreover, it is worth noting the relatively favourable survival outcomes for these LS-associated cancers, which may also influence decision-making [26].

### Other cancer risks

In addition to CRC and gynecologic cancers, elevated risks for urinary and biliary tract cancer were observed in *MSH6* carriers. Comparing these risk with the latest PLSD data [26] suggest similar risk levels but with narrower confidence intervals, affirming the consistency of our findings. However, effective surveillance protocols for UC remain unproven [30].

Regarding breast cancer, recent studies have reported conflicting risks for MSH6 carriers. For example, Dorling et al. [31] identified a modestly increased risk for breast cancer, but the cumulative risk remains significantly lower than some earlier analyses suggested. Our study found a cumulative breast cancer risk for MSH6 carriers of 8% (95% CI, 4.5–14.3) by age 70 and 10.8% (95% CI, 6.2–19) by age 80, which aligns with other prospective analyses showing lower risks than reported in other studies [32, 33].

It is also important to note that in the general population in the Netherlands, approximately 1 in 7 women (14%) will develop breast cancer during their lifetime [13]. In this context, the breast cancer risks we observed for *MSH6* carriers—8% by age 70 and 10.8% by age 80—do not represent a substantial increase over population risk. These findings suggest that breast cancer risk for *MSH6* carriers, it is not markedly higher than the general population risk.

These discrepancies underscore the importance of relying on more robust, unbiased data when counselling *MSH6* carriers about breast cancer risk. Our findings suggest that exaggerated risk estimates should be avoided to ensure accurate counselling and management strategies tailored to individual circumstances.

### Limitations

It is important to acknowledge the limitations of our study. The cohort was primarily clinic-based, which may have led to an overestimation of cancer risk for *MSH6* carriers.

Another limitation of this study is the reliance on self-reported cancer diagnoses from probands or relatives, which were not independently verified. While reports from first-degree relatives are generally considered reliable, discrepancies were observed, particularly for second-degree relatives and certain cancer types, such as endometrial cancer. This highlights the potential for misclassification, which could impact the accuracy of familial cancer assessments. [34].

Furthermore, we did not take into account genetic and environmental risk modifiers. A recent study [5] observed different cancer risks per MMR gene per continent (Europe vs North America vs Australasia), even when the analyses were restricted to one specific *MSH2* variant. This observation stresses the existence of risk modifiers. While our cohort was homogeneously Dutch, which likely reduced the influence of such modifiers, these factors should still be considered in future research for a more comprehensive understanding of cancer risk in *MSH6* carriers.

## Conclusions

In conclusion, our study provides valuable insights for refining clinical guidelines for *MSH6* variant carriers with Lynch syndrome. The data support delaying CRC surveillance to age 35 or 40, without significantly increasing risk. For females, heightened vigilance for EC remains crucial, and risk reducing surgeries should be discussed proactively. Also, there is a need for effective surveillance tools for urinary and biliary tract cancers in LS carriers. Finally, our findings highlight the need for personalized risk estimates and surveillance strategies based on causative gene, age and sex, which can improve patient outcomes while minimizing unnecessary interventions.

## Data Availability

All data produced in the present study are available upon reasonable request to the authors

## Abbreviations

CR: Cumulative Risk
EC: Endometrial Cancer
FDR: First-Degree Relative
HR: Hazard Ratio
LS: Lynch Syndrome
(L)P: (likely) pathogenic
LTR: Lifetime Risk
MMR: Mismatch Repair
NA: Not Applicable
OC: Ovarian Cancer
SD: Standard Deviation
SDR: Second-Degree Relative
UC: urinary tract cancer

## Acknowledgments

We would like to express our gratitude to R.H. van der Eijk, J.E. Nieuwlaat, and M.E. Velthuizen for their assistance in data collection at VUMC, ErasmusMC, and UMCU, respectively. This research received support from the MLDS (Maag Lever Darm Stichting, FP16-06).

